# Co-circulation of SARS-CoV-2 variants B.1.1.7 and P.1

**DOI:** 10.1101/2021.04.06.21254923

**Authors:** Paola Stefanelli, Filippo Trentini, Giorgio Guzzetta, Valentina Marziano, Alessia Mammone, Piero Poletti, Carla Molina Grané, Mattia Manica, Martina del Manso, Xanthi Andrianou, Patrizio Pezzotti, Marco Ajelli, Giovanni Rezza, Silvio Brusaferro, Stefano Merler, COVID-19 National Microbiology Surveillance Study Group

**Affiliations:** Department of Infectious Diseases, Istituto Superiore di Sanità, Rome, Italy; Center for Health Emergencies, Bruno Kessler Foundation, Trento, Italy; Directorate General of Health Prevention, Ministry of Health, Rome, Italy; University of Trento, Italy; European Programme for Intervention Epidemiology Training (EPIET), European Centre for, Disease Prevention and Control (ECDC), Stockholm, Sweden; Cyprus University of Technology, Limassol, Cyprus, Disease Prevention and Control (ECDC), Stockholm, Sweden; Department of Epidemiology and Biostatistics, Indiana University School of Public Health, Bloomington, IN, USA; Laboratory for the Modeling of Biological and Socio-technical Systems, Northeastern University, Boston, MA, USA; Istituto Superiore di Sanità, Rome, Italy; Dept Infectious Diseases, Istituto Superiore di Sanità; Laboratorio di Genetica Molecolare del Centro di Tecnologie Avanzate (CAST) “Università G. d’Annunzio di Chieti”, Chieti; Istituto Zooprofilattico Sperimentale dell’Abruzzo e del Molise “Giuseppe Caporale”, Teramo; A.O.R. San Carlo – Potenza U.O.C di Analisi Chimico Cliniche e Microbiologiche; P.O. Madonna delle Grazie - Matera - U.O.S.D. Laboratorio di Genetica Medica; Laboratorio di Virologia e Microbiologia Azienda Ospedaliera Pugliese-Ciaccio; Francesca Greco UOC di Microbiologia e Virologia PO Annunziata Cosenza; Laboratorio di Genomica e patologia molecolare dell’Università degli studi Magna Graecia di Catanzaro Luigi Atripaldi, AORN Azienda Sanitaria dei Colli, Naple; Istituto Zooprofilattico Sperimentale del Mezzogiorno, Naple; Laboratorio di Virologia – UCO Igiene e Sanità pubblica; Azienda Sanitaria Universitaria Giuliano-Isontina (ASUGI), Trieste; Laboratorio Genomica ed Epigenomica, Area Science Park, Basovizza, Trieste; Stefano Pongolini Istituto Zooprofilattico Sperimentale della Lombardia e dell’Emilia-Romagna – Analisi del Rischio ed Epidemiologia Genomica; Vittorio Sambri DIMES Università di Bologna & U.O.C. Microbiologia AUSL Romagna; U.O.C. Microbiologia AUSL Romagna, Silvia Zannoli U.O.C. Microbiologia AUSL Romagna; Laboratorio di Igiene e Sanità Pubblica, Dipartimento di Medicina e Chirurgia, Università di Parma; Laboratorio di Virologia, Istituto Nazionale Malattie Infettive IRCCS “L.Spallanzani”, Rome; Adrea Orsi Laboratorio di Riferimento Regionale per le Emergenze di Sanità Pubblica (LaRESP); Dipartimento di Scienze Biomediche per la Salute, Università di Milano; Unità Virologia Molecolare, Fondazione IRCCS Policlinico San Matteo, Università di Pavia, Pavia; U.O.C Microbiologia Clinica, Virologia e diagnostica delle Bioemergenze, ASST FBF-Sacco, Milan; S.C. Laboratorio Microbiologia ASST Sette Laghi; ASST Spedali Civili di Brescia; Fondazione IRCCS Ca’ Granda Ospedale Maggiore Policlinico di Milano; Istituto Zooprofilattico Sperimentale della Lombardia e dell’Emilia Romagna-Brescia; Azienda Ospedaliero Universitaria - Ospedali Riuniti Ancona; Massimiliano Scutellà, Ospedale Cardarelli di Campobasso; Laboratorio Aziendale di Microbiologia e Virologia, Azienda Sanitaria dell’Alto Adige; Microbiologia e Virologia - Presidio Ospedaliero Santa Chiara, Trento; Laboratorio di Microbiologia e Virologia ASL Città di Torino; IRCCS-FPO di Candiolo, Giuseppe Ru, Istituto Zooprofilattico Sperimentale del Piemonte; Chironna, Laboratorio di Epidemiologia Molecolare e Sanità Pubblica - AOU Policlinico di Bari; Istituto Zooprofilattico Sperimentale di Puglia e Basilicata per la Puglia; S.C. Microbiologia e Virologia Laboratorio Virologia - AOU di Sassari; Laboratorio Generale (HUB) di analisi chimico cliniche e microbiologia - P.O. Duilio Casula - AOU di Cagliari; Laboratorio di Riferimento Regionale per la Sorveglianza Epidemiologica e Virologica del P.R.O.M.I.S.E. - AOUP “Giaccone” di Palermo; Concetta Ilenia Palermo, Laboratorio di Virologia Clinica - AOUP “V. Emanuele” di Catania - P.O.; Laboratorio di Virologia Laboratorio di Virologia UOC di Microbologia “G. Martino” di Messina; Laboratorio di Diagnostica Molecolare dell’Unità Gestione Centralizzata Laboratori, Messina; CRQ, Sicilia; Stefano Vullo, Stefano Reale, Istituto Zooprofilattico Sperimentale della Sicilia; UOC Microbiologia e Virologia, Azienda Ospedaliera Universitaria Senese Dipartimento Biotecnologie Mediche, Università degli Studi di Siena; SOD Microbiologia e Virologia Azienda Ospedaliera Universitaria Careggi; UOC Virologia Azienda Ospedaliero-Universitaria Pisana, Pisa; Microbiology and Clinical Microbiology, Department of Medicine and Surgery, University of Perugia; Istituto Zooprofilattico Sperimentale delle Regioni Umbria e Marche; Laboratorio Analisi Cliniche dell’Ospedale Parini di Aosta; Istituto Zooprofilattico Sperimentale delle Venezie; U.O.C. Microbiologia-Virologia-AULSS2 La Marca - P.O. Treviso

**Author notes:** corresponding author **Corresponding author: Stefano Merler**. contributed equally (Paola Stefanelli, Filippo Trentini). senior authors (Marco Ajelli, Giovanni Rezza, Silvio Brusaferro, Stefano Merler).

## Abstract

SARS-CoV-2 variants of concern (B.1.1.7, P.1 and B.1.351) have emerged in different continents of the world. To date, little information is available on their ecological interactions. Based on two genomic surveillance surveys conducted on February 18 and March 18, 2021 across the whole Italian territory and covering over 3,000 clinical samples, we found significant co-circulation of B.1.1.7 and P.1. We showed that B.1.1.7 was already dominant on February 18 in a majority of regions/autonomous provinces (national prevalence 54%) and almost completely replaced historical lineages by March 18 (dominant in all regions/autonomous provinces, national prevalence 86%). At the same time, we found a substantial proportion of cases of the P.1 lineage on February 18, almost exclusively in Central Italy (with an overall prevalence in the macro-area of 18%), which remained at similar values on March 18, suggesting the inability by this lineage to outcompete B.1.1.7. Only 9 cases from variant B.1.351 were identified in the two surveys. At the national level, we estimated a mean relative transmissibility of B.1.1.7 (compared to historical lineages) ranging between 1.55 and 1.57 (with confidence intervals between 1.45 and 1.66). The relative transmissibility of P.1 estimated at the national level varied according to the assumed degree of cross-protection granted by infection with other lineages and ranged from 1.12 (95%CI 1.03-1.23) in the case of complete immune evasion by P.1 to 1.39 (95%CI 1.26-1.56) in the case of complete cross-protection. These observations may have important consequences on the assessment of future pandemic scenarios.

## Introduction

Since the end of 2020, multiple SARS-CoV-2 variants have emerged across the globe. Some of these are particularly concerning as their biological characteristics allowed them to outcompete and rapidly replace historical lineages in countries where they likely emerged, and to spread rapidly to many other countries. Variant B.1.1.7 was first detected in the United Kingdom from samples of September 2020 and was dominant throughout the country by early 2021 [1, 2]; it has spread in most of Europe [3] and it has been reported in a majority of world countries [4, 5, 6]. Variant P.1 was first reported in Japan among travelers returning from Brazil [7]. It was later found in almost half of cases from December 2020 in Manaus, Brazil [8] where, despite a very high estimated seroprevalence against historical lineages [9], a large upsurge of infections occurred throughout January 2021 [10]. B.1.351 was first detected in South Africa, where it is dominant since late November [11]. The epidemiological success of these variants relies on evolutionary advantages such as increased transmissibility [1, 2, 12] and their ability (demonstrated for P.1 and B.1.351) to significantly reduce neutralization in convalescent and post-vaccination sera [13, 14, 15, 16, 17], likely resulting in reinfections through immune escape [18, 19, 20, 21]. Besides their greater ability to spread, requiring more restrictive physical distancing measures to mitigate epidemics, these variants cause additional concern due to potential increased morbidity [22] and mortality [23] (currently evaluated for B.1.1.7 only), as well as their potential impact on current vaccine effectiveness [24, 25]. Because variants seem to have emerged in disjoint geographical areas, little is known as yet about their ecological interactions. Here, we provide the first observation of significant co-circulation of variants B.1.1.7 and P.1 using data from genomic surveillance surveys in Italy. We use a mathematical model to estimate their relative transmissibility under different assumptions on the degree cross-protection.

## Methods

### Survey methodology

Two surveys coordinated by the Italian National Institute of Health (Istituto Superiore di Sanità), in collaboration with the Ministry of Health and the Regions/Autonomous Provinces (AP), aimed at estimating the prevalence of the B.1.1.7, P.1, and B.1.351 lineages, were conducted on February 18, 2021 and March 18, 2021. Istituto Superiore di Sanità obtained an ethical approval for sequencing SARS-CoV-2 genomes on clinical samples (ref. PRE BIO CE n.26259, July 29, 2020).

The surveys involved all of the 19 Regions and 2 Autonomous Provinces (AP) of Italy. Random samples of cases, diagnosed on February 18 and March 18 with a real-time reverse transcription polymerase chain reaction (RT-PCR) having cycle threshold (CT) < 28, were analyzed in 101 and 126 laboratories, respectively, distributed across the national territory.

Samples were uniformly distributed across 5 macro-areas, defined according to the Eurostat classification: North-East, North-West, Center, South and Islands. The sample size was calculated to have the statistical power to detect a prevalence of 1%, with 0,8% error within each macro-area, based on the number of cases notified on the day preceding each of the two surveys. The collected samples were sequenced according to the local laboratory policy by either of the following techniques: i) sequencing the entire S-gene by Sanger technology, ii) sequencing part of the S-gene with the identification of all mutations/deletions attributable to the three variants, or iii) sequencing the whole genome by Next Generation Sequencing. In the second survey, one region (Marche) pre-screened 54 of the 65 RT-PCR positive samples using an in-house test that detects both H69-V70 and Y144 aminoacid deletions, which are specific for B.1.1.7; as a result, 46 cases positive to the in-house test were considered B.1.1.7 without sequencing. The 8 cases negative to the test, plus the 11 cases that were not pre-screened were sequenced.

The point prevalence of the three lineages in each survey was computed as the fraction of infections attributable to each lineage among sequenced samples, and corresponding binomial 95% confidence intervals are provided. In the second survey, data from Marche were excluded from the analysis.

### SARS-CoV-2 three-strain transmission model

We adopted a three-strain Susceptible – Infectious – Recovered (SIR) mathematical model to simulate co-circulation of historical lineages of SARS-CoV-2 (”wildtype”) and variants of concern B.1.1.7 and P.1. We did not consider B.1.351 based on results from the surveys finding little or no circulation of this lineage (see Results). We assumed that a previous infection with the wildtype provides complete protection against variant B.1.1.7 [26] and that infection with either the wildtype or B.1.1.7 confers the same degree of cross-protection *χ* against P.1. In addition, we assumed that the transmissibility of variants B.1.1.7 and P.1 are scaled with respect to the wildtype transmissibility by a lineage-specific factor representing their relative transmissibility (k1 for B.1.1.7 and k2 for P.1). The three strains are assumed to have identical generation time (set at 6.6 days to reflect the serial interval estimated for SARS-CoV-2 in Italy [27]). The model is initialized on January 15, 2021, assuming that an unknown fraction f1 and f2 of all infections at that date belongs to the B.1.1.7 and P.1 variant, respectively (see Appendix for full model details). Unknown model parameters (namely, the transmissibility of the wildtype strain *β*, the relative transmissibility parameters of B.1.1.7 and P.1, k1 and k2, and their respective initial prevalence, f1 and f2) were estimated by calibrating the model against prevalence data from the two surveys (Table 1 and 2), using a Markov Chain Monte Carlo (MCMC) approach. The likelihood was defined as:

**Table 1.** Results of the first survey (February 18, 2021) across the 21 participating regions/AP.

| MACRO-AREA | REGION | LABS | RT-PCR<br>POSITIVE | SEQUENCED<br>SAMPLES | ANALYZED<br>SAMPLES | CONFIRMED CASES |  |  | POINT PREVALENCE |  |  |
| --- | --- | --- | --- | --- | --- | --- | --- | --- | --- | --- | --- |
|  |  |  |  |  |  | B.1.1.7 | P.1 | B.1.1.135 | B.1.1.7 | P.1 | B.1.1.135 |
| ISLANDS | SARDINIA | 6 | 38 | 25 | 12 | 9 | 0 | 0 | 75.0%<br>(42.8-94.5) | 0.0%<br>(0-26.5) | 0.0%<br>(0-26.5) |
|  | SICILY | 5 | 268 | 63 | 58 | 32 | 0 | 1 | 55.2%<br>(41.5-68.3) | 0.0%<br>(0-6.2) | 1.7%<br>(0-9.2) |
|  | TOTAL ISLANDS | 11 | 306 | 88 | 70 | 41 | 0 | 1 | 58.6%<br>(46.2-70.2) | 0.0%<br>(0-5.1) | 1.4%<br>(0.5-7.7) |
| SOUTH | ABRUZZO | 2 | 374 | 61 | 61 | 31 | 0 | 0 | 50.8%<br>(37.7-63.9) | 0.0%<br>(0-5.9) | 0.0%<br>(0-5.9) |
|  | APULIA | 7 | 59 | 59 | 59 | 28 | 0 | 0 | 47.5%<br>(34.3-60.9) | 0.0%<br>(0-6.1) | 0.0%<br>(0-6.1) |
|  | BASILICATA | 5 | 7 | 7 | 5 | 1 | 0 | 0 | 20.0%<br>(0.5-71.6) | 0.0%<br>(0-52.2) | 0.0%<br>(0-52.2) |
|  | CALABRIA | 3 | 166 | 11 | 11 | 1 | 0 | 0 | 9.1%<br>(0.2-41.3) | 0.0%<br>(0-28.5) | 0.0%<br>(0-28.5) |
|  | CAMPANIA | 2 | 366 | 86 | 86 | 51 | 2 | 0 | 59.3%<br>(48.2-69.8) | 2.3%<br>(0.3-8.1) | 0.0%<br>(0-4.2) |
|  | MOLISE | 1 | 114 | 15 | 15 | 14 | 0 | 0 | 93.3%<br>(68.0-99.8) | 0.0%<br>(0-21.8) | 0.0%<br>(0-21.8) |
|  | TOTAL SOUTH | 20 | 1086 | 239 | 237 | 126 | 2 | 0 | 53.2%<br>(46.6-59.7) | 0.8%<br>(0.1-3.0) | 0.0%<br>(0-1.5) |
| CENTER | LAZIO | 5 | 169 | 169 | 144 | 49 | 19 | 0 | 34.0%<br>(26.3-42.4) | 13.2%<br>(8.1-19.8) | 0.0%<br>(0-2.5) |
|  | MARCHE | 8 | 38 | 38 | 38 | 22 | 3 | 0 | 57.9%<br>(40.8-73.7) | 7.9%<br>(1.7-21.4) | 0.0%<br>(0-9.3) |
|  | TUSCANY | 3 | 88 | 80 | 80 | 43 | 19 | 0 | 53.8%<br>(42.2-65.0) | 23.8%<br>(14.9-34.6) | 0.0%<br>(0-4.5) |
|  | UMBRIA | 4 | 247 | 48 | 47 | 24 | 17 | 0 | 51.1%<br>(36.1-65.9) | 36.2%<br>(22.7-51.5) | 0.0%<br>(0-7.5) |
|  | TOTAL CENTER | 20 | 542 | 335 | 309 | 138 | 58 | 0 | 44.7%<br>(39.0-50.4) | 18.8%<br>(14.6-23.6) | 0.0%<br>(0-1.2) |
| NORTH-EAST | AP BOLZANO | 1 | 320 | 70 | 70 | 40 | 0 | 2 | 57.1%<br>(44.7-68.9) | 0.0%<br>(0-5.1) | 2.9%<br>(0.3-9.9) |
|  | AP TRENTO | 1 | 20 | 20 | 14 | 2 | 0 | 0 | 14.3%<br>(1.8-42.8) | 0.0%<br>(0-23.2) | 0.0%<br>(0-23.2) |
|  | EMILIA-ROMAGNA | 2 | 99 | 99 | 99 | 57 | 2 | 0 | 57.6%<br>(47.2-67.4) | 2.0%<br>(0.2-7.1) | 0.0%<br>(0-3.7) |
|  | FRIULI VENEZIA GIULIA | 4 | 133 | 28 | 27 | 8 | 0 | 0 | 29.6%<br>(13.7-50.2) | 0.0%<br>(0-12.8) | 0.0%<br>(0-12.8) |
|  | VENETO | 12 | 92 | 92 | 92 | 52 | 0 | 0 | 56.5%<br>(45.8-66.8) | 0.0%<br>(0-3.9) | 0.0%<br>(0-3.9) |
|  | TOTAL NORTH-EAST | 20 | 664 | 309 | 302 | 159 | 2 | 2 | 52.6%<br>(46.9-58.4) | 0.7%<br>(0.1-2.4) | 0.7%<br>(0.1-2.4) |
| NORTH-WEST | AOSTA VALLEY | 1 | 1 | 1 | 1 | 0 | 0 | 0 | 0.0%<br>(0-97.5) | 0.0%<br>(0-97.5) | 0.0%<br>(0-97.5) |
|  | LIGURIA | 6 | 227 | 22 | 22 | 16 | 0 | 0 | 72.7%<br>(49.8-89.3) | 0.0%<br>(0-15.4) | 0.0%<br>(0-15.4) |
|  | LOMBARDY | 9 | 213 | 213 | 213 | 137 | 0 | 3 | 64.3%<br>(57.5-70.7) | 0.0%<br>(0-1.7) | 1.4%<br>(0.3-4.1) |
|  | PIEDMONT | 14 | 93 | 89 | 85 | 41 | 0 | 0 | 48.2%<br>(37.3-59.3) | 0.0%<br>(0-4.2) | 0.0%<br>(0-4.2) |
|  | TOTAL NORTH-WEST | 30 | 534 | 325 | 321 | 194 | 0 | 3 | 60.4%<br>(54.9-65.8) | 0.0%<br>(0-1.1) | 0.9%<br>(0.2-2.7) |
| TOTAL ITALY |  | 101 | 3132 | 1296 | 1239 | 658 | 62 | 6 | 53.1%<br>(50.3-55.9) | 5.0%<br>(3.9-6.4) | 0.5%<br>(0.2-1.1) |

**Table 2.** Results from the second survey (March 18, 2021) across the 21 participating regions/AP.

| MACRO-AREA | REGION | LABS | RT-PCR<br>POSITIVE | SEQUENCED<br>SAMPLES | ANALYZED<br>SAMPLES | CONFIRMED CASES |  |  | POINT PREVALENCE |  |  |
| --- | --- | --- | --- | --- | --- | --- | --- | --- | --- | --- | --- |
|  |  |  |  |  |  | B.1.1.7 | P.1 | B.1.1.135 | B.1.1.7 | P.1 | B.1.1.135 |
| ISLANDS | SARDINIA | 6 | 85 | 21 | 21 | 18 | 0 | 1 | 85.7%<br>(63.7-97) | 0.0%<br>(0-16.1) | 4.8%<br>(0.1-23.8) |
|  | SICILY | 5 | 632 | 132 | 129 | 97 | 3 | 0 | 75.2%<br>(66.8-82.4) | 2.3%<br>(0.5-6.6) | 0.0%<br>(0-2.8) |
|  | TOTAL ISLANDS | 11 | 717 | 153 | 150 | 115 | 3 | 1 | 76.5%<br>(69.1-83.2) | 2.0%<br>(0.4-5.7) | 0.7%<br>(0.0-3.7) |
| SOUTH | ABRUZZO | 2 | 293 | 87 | 80 | 66 | 4 | 0 | 82.5%<br>(72.4-90.1) | 5.0%<br>(1.4-12.3) | 0.0%<br>(0-4.5) |
|  | APULIA | 11 | 126 | 126 | 126 | 117 | 0 | 0 | 92.9%<br>(86.9-96.7) | 0.0%<br>(0-2.9) | 0.0%<br>(0-2.9) |
|  | BASILICATA | 6 | 62 | 27 | 20 | 13 | 0 | 0 | 65.0%<br>(40.8-84.6) | 0.0%<br>(0-16.8) | 0.0%<br>(0-16.8) |
|  | CALABRIA | 4 | 404 | 26 | 26 | 22 | 0 | 0 | 84.6%<br>(65.1-95.6) | 0.0%<br>(0-13.2) | 0.0%<br>(0-13.2) |
|  | CAMPANIA | 3 | 1400 | 261 | 261 | 232 | 4 | 0 | 88.9%<br>(84.4-92.4) | 1.5%<br>(0.4-3.9) | 0.0%<br>(0-1.4) |
|  | MOLISE | 1 | 63 | 16 | 16 | 13 | 2 | 0 | 81.2%<br>(54.4-96) | 12.5%<br>(1.6-38.3) | 0.0%<br>(0-20.6) |
|  | TOTAL SOUTH | 27 | 2348 | 543 | 529 | 463 | 10 | 0 | 87.5%<br>(84.4-90.2) | 1.9%<br>(0.9-3.4) | 0%<br>(0.0-0.7) |
| CENTER | LAZIO | 11 | 214 | 205 | 205 | 161 | 42 | 0 | 78.5%<br>(72.3-84) | 20.5%<br>(15.2-26.7) | 0.0%<br>(0-1.8) |
|  | MARCHE* | 11 | 65 | - | - | - | - | - | - | - | - |
|  | TUSCANY | 3 | 144 | 103 | 99 | 85 | 10 | 0 | 85.9%<br>(77.4-92) | 10.1%<br>(5-17.8) | 0.0%<br>(0-3.7) |
|  | UMBRIA | 5 | 80 | 26 | 25 | 16 | 8 | 0 | 64%<br>(42.5-82) | 32.0%<br>(14.9-53.5) | 0.0%<br>(0-13.7) |
|  | TOTAL CENTER # | 30 | 503 | 334 | 329 | 262 | 60 | 0 | 79.6%<br>(74.9-83.9) | 18.2%<br>(14.2-22.8) | 0%<br>(0.0-1.1) |
| NORTH-EAST | AP BOLZANO | 1 | 69 | 15 | 15 | 12 | 0 | 0 | 80.0%<br>(51.9-95.7) | 0.0%<br>(0-21.8) | 0.0%<br>(0-21.8) |
|  | AP TRENTO | 1 | 16 | 16 | 16 | 16 | 0 | 0 | 100.0%<br>(79.4-100) | 0.0%<br>(0-20.6) | 0.0%<br>(0-20.6) |
|  | EMILIA-ROMAGNA | 2 | 175 | 175 | 175 | 154 | 13 | 0 | 88.0%<br>(82.2-92.4) | 7.4%<br>(4.0-12.4) | 0.0%<br>(0-2.1) |
|  | FRIULI VENEZIA GIULIA | 7 | 126 | 55 | 55 | 49 | 0 | 0 | 89.1%<br>(77.8-95.9) | 0.0%<br>(0-6.5) | 0.0%<br>(0-6.5) |
|  | VENETO | 13 | 156 | 156 | 156 | 138 | 2 | 1 | 88.5%<br>(82.4-93) | 1.3%<br>(0.2-4.6) | 0.6%<br>(0-3.5) |
|  | TOTAL NORTH-EAST | 24 | 536 | 417 | 417 | 369 | 15 | 1 | 88.5%<br>(85.0-91.4) | 3.6%<br>(2.0-5.9) | 0.2%<br>(0.0-1.3) |
| NORTH-WEST | AOSTA VALLEY | 1 | 32 | 2 | 2 | 2 | 0 | 0 | 100.0%<br>(15.8-100) | 0.0%<br>(0-84.2) | 0.0%<br>(0-84.2) |
|  | LIGURIA | 8 | 179 | 22 | 22 | 14 | 3 | 0 | 63.6%<br>(40.7-82.8) | 13.6%<br>(2.9-34.9) | 0.0%<br>(0-15.4) |
|  | LOMBARDY | 12 | 314 | 314 | 312 | 278 | 0 | 1 | 89.1%<br>(85.1-92.3) | 0.0%<br>(0-1.2) | 0.3%<br>(0-1.8) |
|  | PIEDMONT | 16 | 155 | 153 | 153 | 138 | 1 | 0 | 90.2%<br>(84.3-94.4) | 0.7%<br>(0-3.6) | 0.0%<br>(0-2.4) |
|  | TOTAL NORTH-WEST | 37 | 680 | 491 | 489 | 432 | 4 | 1 | 88.3%<br>(85.2-91.1) | 0.8%<br>(0.2-2.1) | 0.2%<br>(0.0-1.1) |
| TOTAL ITALY # |  | 129 | 4790 | 1938 | 1914 | 1641 | 92 | 3 | 85.7%<br>(84.1-87.3) | 4.8%<br>(3.9-5.9) | 0.2%<br>(0.0-0.5) |

\* Results for Marche are not reported as the regional laboratory followed a different experimental design (see Methods).

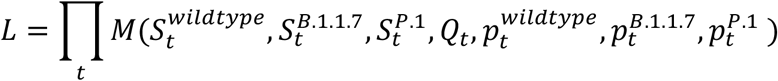

Where *M*(·) is the multinomial probability density distribution; t is the date of the survey; 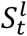 is the number of cases observed at date t for lineage *l*;, Q_t_ is the total number of sequenced cases at date t; and 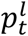 is the model-estimated fraction of infections of lineage *l* over the total at date t. We assigned L=0 to simulations for which the model’s mean squared error on the observed daily hospital admissions between January 18 and March 18, 2021 exceeded 1.5 times the variance of observations, in order to reproduce the observed epidemiological temporal trends.

The model was run on 4 different geographical aggregations of regional data, i.e., the national level and the Center, North-East, and South macro-areas (see Table 1 and 2). Macro-areas North-West and Islands did not have a sufficient number of P.1 cases. Data from Marche were excluded due to heterogeneity in data collection.

## Results

### Prevalence of variants of concern in Italy, February 18 and March 18

During the first genomic survey conducted on February 18, 1,296 samples were sequenced, of which 57 (4%) were discarded for the analysis due to insufficient sequencing coverage of the genome. Among the remaining 1,239 samples, 658 infections were attributable to the SARS-CoV-2 lineage B.1.1.7, 62 to lineage P.1 and 6 to lineage B.1.351, for a national prevalence of 53.1% (95%CI: 50.3-55.9), 5.0% (96%CI: 3.9-6.4) and 0.5% (95%CI: 0.2-1.1), respectively. B.1.1.7 was found in 20 of 21 regions/AP, P.1 in 6, and B.1.351 in 3, as shown in Figure 1 and Table 1. The prevalence of B.1.1.7 was highest in the North-West macro-area (60.4%, 95%CI: 54.9-65.8%) and lowest in the Center (44.7%, 95%CI: 39.0-50.4), while P.1 was almost exclusively concentrated in the Center (mean prevalence 18.8%, 95%CI: 14.6-23.6, as opposed to 1% or less elsewhere; see Table 1 and Figure 1). B.1.351 was identified only in Lombardy (3 cases), in the AP Bolzano (2 cases) and in Sicily (1 case).

**Figure 1.**
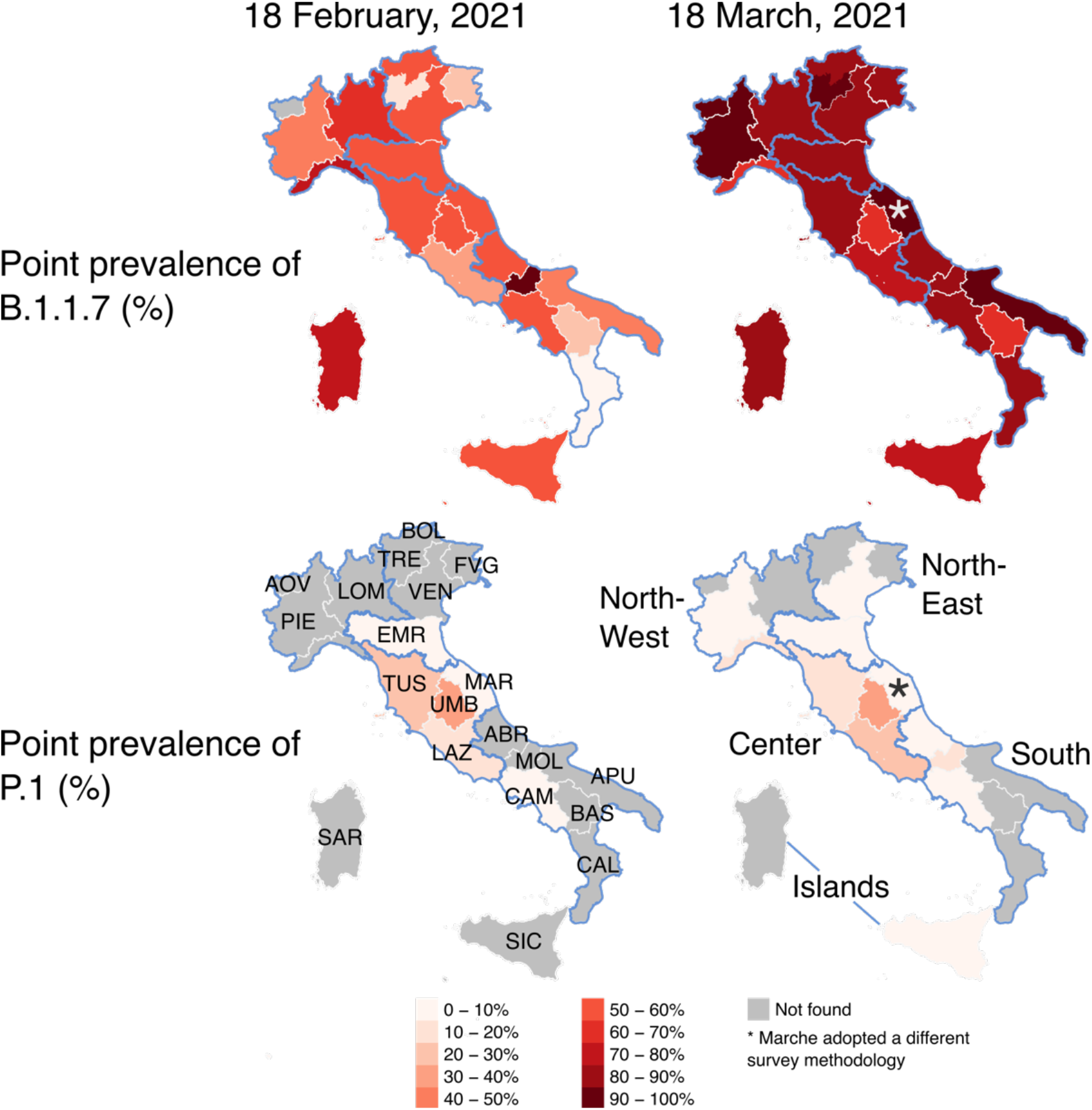
Geographical distribution of variants of concern. Point prevalence of lineages B.1.1.7 (top row) and P.1 (bottom row) by region/autonomous province of Italy as obtained from the national surveys conducted on February 18 (left) and March 18 (right), 2021. Abbreviations of region names are reported in the bottom-left map. Borders of mainland macro-areas are highlighted in blue and the names of macro-areas are reported in the bottom-right map.

During the second genomic survey conducted on March 18, 1,938 samples were sequenced (not including data from Marche), of which 24 (1%) were discarded for the analysis due to lack of enough coverage of the genome sequencing (Table 2). Among the remaining 1,914 samples, 1,641 infections were attributable to the SARS-CoV-2 lineage B.1.1.7, 92 to lineage P.1 and 3 to lineage B.1.351, for a national prevalence of 85.7% (95%CI: 84.1-87.3), 4.8% (95%CI: 3.9-5.9) and 0.2% (95%CI: 0.0-0.4%), respectively. B.1.1.7 was found in all 21 regions/AP, P.1 in 12, and B.1.351 in 3, as shown in Figure 1 and Table 2. According to the survey conducted on March 18, B.1.1.7 has become dominant in all Italian regions, with regional prevalence estimates ranging from 63.6% to 100% (Figure 1 and Table 2). Regional prevalence estimates of P.1 range from 0% to 32%; the highest prevalence estimates are still obtained for central regions (Figure 1), however, this lineage has been detected in six additional regions compared to February 18. B.1.351 was identified only in Lombardy, Sardinia and Veneto (1 case each).

### Relative transmissibility of B.1.1.7 and P.1

The model was able to fit the epidemiological trends on hospital admissions and the estimated prevalence of B.1.1.7 and P.1 in all geographical aggregations and independently of the assumed degree of cross-protection. Figure 2 shows results for Italy when assuming no cross-protection or complete cross-protection. Independently on the geographical aggregation and on the assumed degree of cross-protections, we found a robust mean estimate for the relative transmissibility of B.1.1.7, ranging between 1.48 and 1.73, with confidence intervals ranging between 1.31 and 1.97 (Figure 3). Considering the national aggregation, estimates varied between 1.55 and 1.57 with confidence intervals ranging between 1.45 and 1.66. The estimated relative transmissibility of P.1 was systematically lower. For the national aggregation and for the three regions of the macro-area “Center” (Lazio, Tuscany and Umbria) where the observed prevalence of P.1 was higher, we estimate a relative transmissibility of 1.12-1.24 under the assumption of no cross-protection (range of 95% CI: 1.03 −1.42), growing linearly for increasing values of cross-protection, up to 1.39-1.46 (range of 95% CI: 1.26-1.63) under the assumption of complete cross-protection. Results for macro-areas North-East and South reproduce a similar pattern, although estimates are more variable due to the limited number of reported P.1 cases.

**Figure 2.**
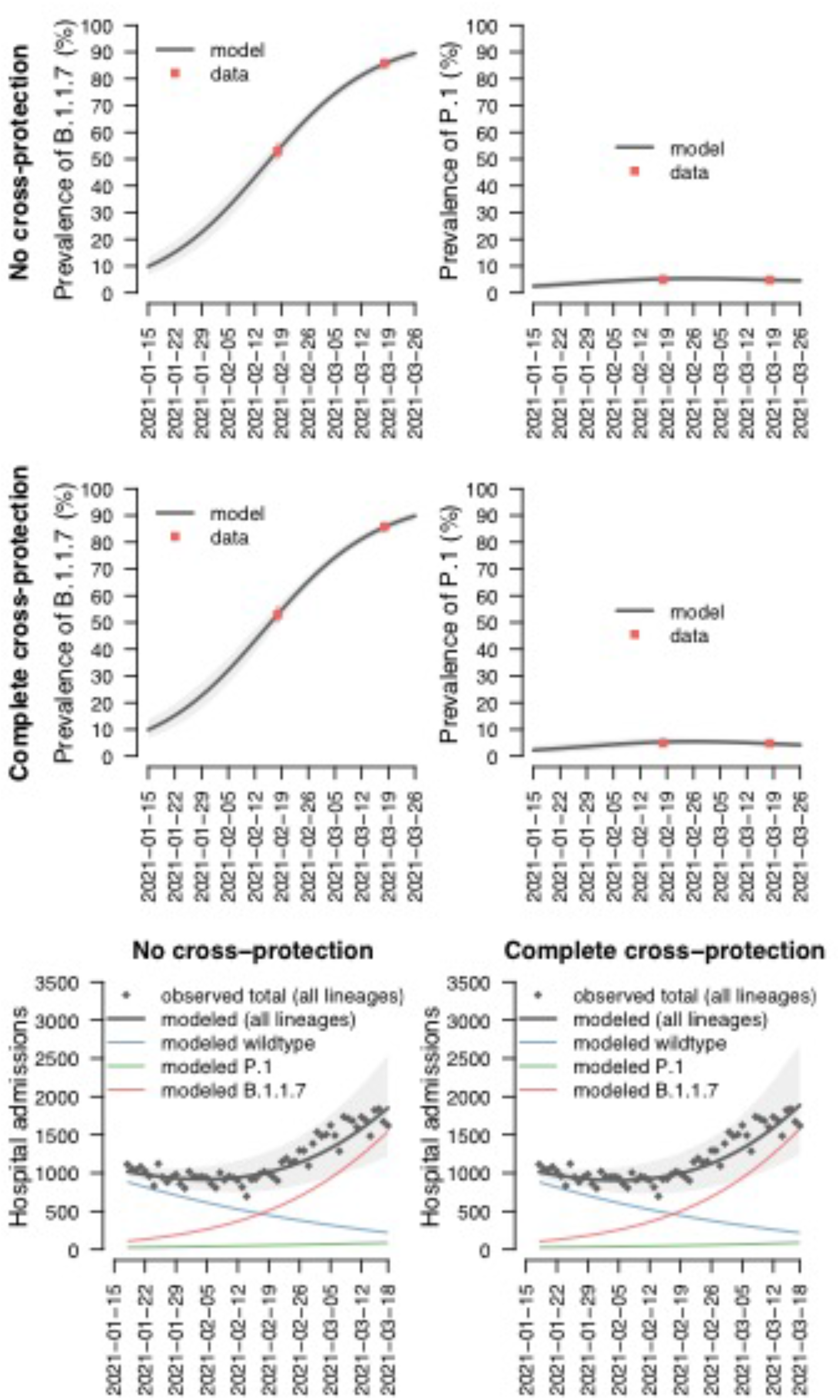
Model fits. Top row: model-estimated (solid black lines: mean values; shaded areas: 95% CI) and observed (red points: mean values; red lines: 95%CI) prevalence of B.1.1.7 (left) and P.1 (right) when assuming no cross-protection between wildtype or B.1.1.7 and P.1. Middle row: as top row but assuming complete cross-protection among all lineages. Bottom row: model-estimated and observed hospital admissions over time. Black lines represent mean values of model-estimated overall daily hospitalizations, shaded areas indicate 95%CI. Colored lines indicate mean values of model-estimated daily hospitalizations attributable to wildtype (blue), B.1.1.7 (red) and P.1 (green) lineages. Black points indicate observed daily hospitalizations.

**Figure 3.**
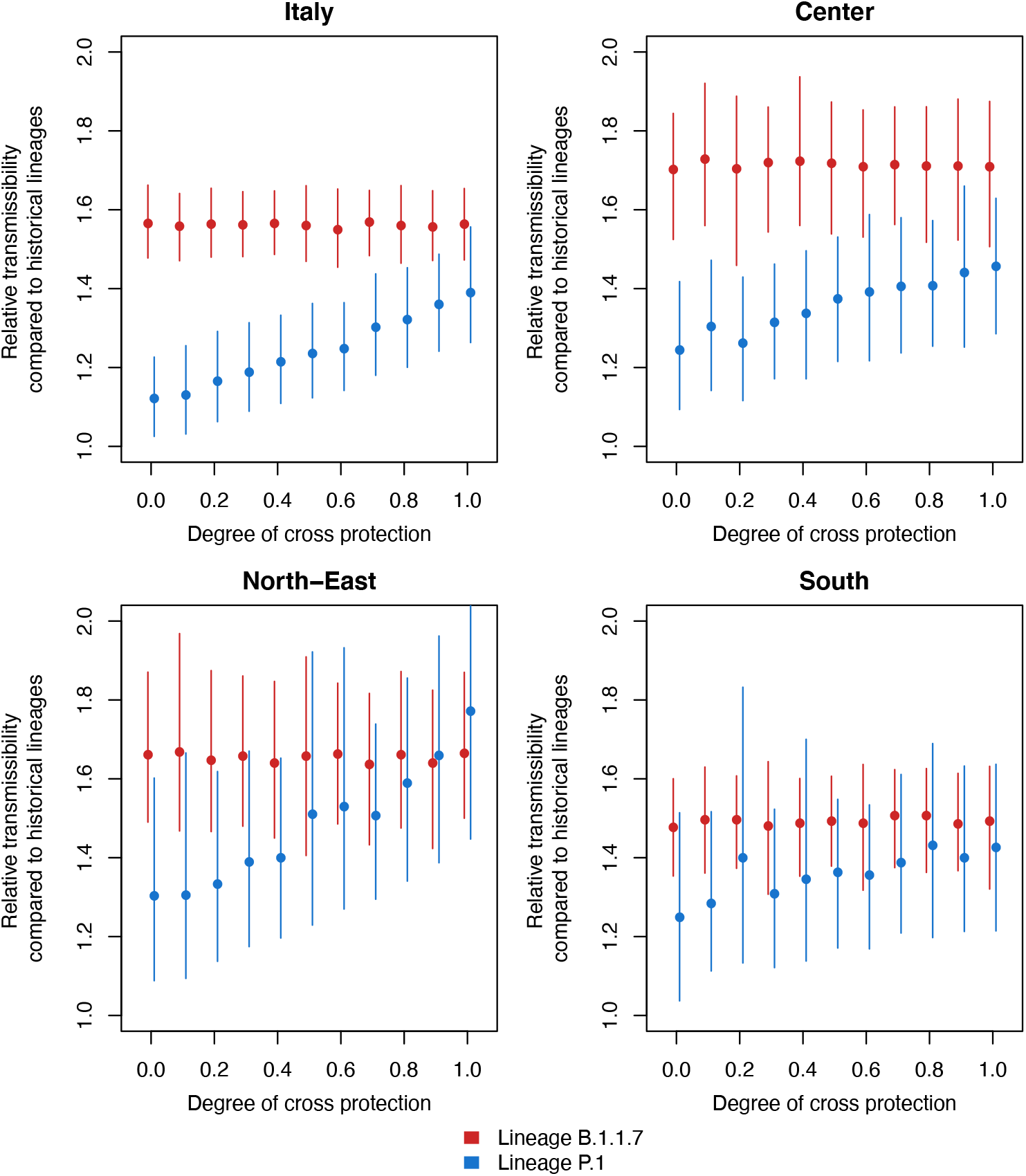
Estimates of the relative transmissibility of B.1.1.7 and P.1. Estimates are provided for different assumed values on the degree of cross protection (0: no cross-protection; 1: complete cross-protection) conferred by previous infection with the wildtype or B.1.1.7 against reinfection with P.1. Points indicate the mean value of the estimated relative transmissibility of B.1.1.7 (red) and P.1 (blue) lineages; lines indicate 95%CI. Panels represent four geographical aggregations for which the analysis was possible (Italy as whole, Center, North-East and South) given the presence of the P.1 lineage. Data from Marche were excluded from the estimate for the national and macro-area “Center” due to heterogeneity in data collection.

## Discussion

Based on two genomic surveillance surveys conducted over the whole Italian territory on February 18 and March 18, 2021, we reported the first observations on significant co-circulation of the B.1.1.7 and P.1 variants of SARS-CoV-2 in Italy. We showed that B.1.1.7 was already dominant on February 18 in a majority of regions/AP (national prevalence 54%) and almost completely replaced historical lineages by March 18 (dominant in all regions/AP, national prevalence 87%). At the same time, we found a substantial proportion of cases of the P.1 lineage on February 18, almost exclusively in regions of Central Italy (Lazio, Tuscany, Umbria and Marche, with an overall prevalence of 18%). P.1 was also identified in samples from Campania and Emilia Romagna, both with prevalence below 3%. The prevalence of P.1 remained similar on March 18, suggesting the inability by this lineage to outcompete B.1.1.7. However, on March 18 lineage P.1 was identified in cases from six additional regions in Northern (Piedmont, Veneto, Liguria) and Southern Italy (Abruzzo, Molise, Sicily). We found only 6 cases from the B.1.351 lineage among the 1239 analyzed samples on February 18, and only 3 of 1908 on March 18. Compared to historical lineages, we estimated a mean relative transmissibility of B.1.1.7 ranging between 1.55 and 1.57 (with confidence intervals between 1.45 and 1.66) in Italy. These values are consistent with available estimates from UK [1, 2] and France [12].

The estimated relative transmissibility of P.1 (compared to historical lineages) varied according to different assumptions on the degree of cross-protection granted by previous infection with historical lineages or B.1.1.7: the estimate at the national level ranged from 1.12 (95%CI 1.03-1.23) in the case of complete immune evasion by P.1 to 1.39 (95%CI 1.26-1.56) in the case of complete cross-protection. Previous estimates on the relative transmissibility of P.1, provided from a study in Manaus, Brazil, where the variant rapidly replaced historical lineages, were very broad (between 1.03 and 2.87), with an estimate of cross-protection between 12 and 90%. The true degree of cross-protection between B.1.1.7 and P.1 is likely critical for the coexistence of B.1.1.7 or P.1, and a key role will be played by the effectiveness of licensed vaccines against the two strains. Importantly, the slight decrease of P.1 prevalence over one month occurred under a condition of strict mitigation measures; if P.1 can at least partially escape immunity from B.1.1.7 and existing vaccines, this may pose challenges towards releasing physical distancing measures as population immunity grows. Furthermore, if some degree of cross-protection exists, a higher proportion of asymptomatic infections may be observed, posing challenges to surveillance and control.

We acknowledge a number of limitations for this study. The sample size was calculated to have the statistical power to detect different lineages at the macro-area level. As such, regional estimates of prevalence should be taken with caution due to the small number of sequenced samples. Samples were randomly selected for sequencing among cases diagnosed by the labs, but some degree of correlation between them (e.g., cases belonging to an over-represented cluster on that day) cannot be completely excluded, especially in regions with lower sample sizes. Due to the different laboratory methodology of one region in the second survey, those data were excluded from the computation of the national prevalence of the variants. Possible biases in the estimate are expected to be minimal, since cases from this region represented only about 1.5% of the total.

For what concerns model estimates on transmissibility, we could not take into account possible differences across strains in the duration of viral shedding [28], age-specific susceptibility or transmissibility [29], the proportion of asymptomatic individuals and their relative transmissibility [29], or other major determinants of the transmission dynamics of SARS-CoV-2, due to the lack of available data on the variants. Similarly, we could not distinguish the potential lineage-specific impact of existing mitigation measures in Italy, nor factor in the potential impact of vaccinations. These factors will likely shape the outcome of the epidemiological competition across strains.

Despite these limitations, we provide evidence that the P.1 lineage was not able to outcompete B.1.1.7 in Italy in over a month of co-circulation, under existing mitigation measures. We suggest that this may be due to a lower transmissibility of P.1, independently of its ability to re-infect individuals previously infected by historical or wildtype strains.

## Data Availability

All data for this study are reported in the manuscript tables or are publicly available at https://www.epicentro.iss.it/coronavirus/open-data/covid_19-iss.xlsx.

## Acknowledgements

The Authors thank Francesco Maraglino, Ministry of Health, for supporting coordination activities.

